# Balanced crystalloids compared to normal saline for fluid therapy in critically ill adult patients: Systematic review and meta-analysis protocol

**DOI:** 10.1101/2021.08.24.21262515

**Authors:** Naomi E Hammond, Fernando G Zampieri, Tessa Garside, Derek Adigbli, Gian Luca Di Tanna, Alexandre B Cavalcanti, Flavia R Machado, Sharon Micallef, John Myburgh, Mahesh Ramanan, Todd W Rice, Matthew Semler, Paul J Young, Balasubramanian Venkatesh, Simon Finfer, Anthony Delaney

## Abstract

**Introduction:** The choice of intravenous fluid for fluid therapy in critically ill adult patients remains a matter of debate. Currently, crystalloids are used more often than colloids, with ongoing controversy over the relative efficacy and safety of buffered salt solutions (BSS) versus normal saline (0.9% sodium chloride). In 2021 two large pragmatic trials enrolling critically ill patients will add substantial new data to address this controversy. We will conduct a systematic review and meta-analysis of randomised controlled trials (RCTs) that will include the data from these two trials to provide clinicians with the most up to date evidence and robust evidence to guide their choice of crystalloid fluids.

**Methods and analysis:** We will include RCTs that compare the effect of buffered salt solutions to normal saline for fluid resuscitation and/or fluid therapy in critically ill adults, on all-cause mortality and other patient centred outcomes. We will perform a search that includes the electronic databases MEDLINE and EMBASE, and clinical trial registries. Two reviewers will independently screen titles and abstracts, perform full article reviews and extract study data, with discrepancies resolved by a third reviewer. We will report study characteristics and assess risk of bias using the Cochrane Risk-of-Bias tool. We will perform Hartung-Knapp-Sidik-Jonkman random-effects aggregate data meta-analysis whenever it is feasible to do so. We will evaluate overall certainty of evidence using the Grading of Recommendations Assessment, Development, and Evaluation (GRADE) framework.

**Ethics and dissemination:** This systematic review and meta-analysis does not require ethical approval as it does not involve primary data collection. We will publish our results in a peer-reviewed scientific journal and present them at national and international scientific conferences.

**PROSPERO registration number:** CRD42021243399

**Strengths and limitations of this review [box]:** This systematic review will provide up-to-date evidence to answer the focused clinical question: In adult patients who are critically ill, does administering balanced crystalloid solutions for fluid therapy reduce mortality and other patient-centered outcomes, compared with administering 0.9% sodium chloride?

We will conduct a systematic review according to the Preferred Reporting Items for Systematic Review and Meta-Analysis Protocols guidelines, searching three electronic databases, clinical trial registries and published conference abstracts, with two independent reviewers evaluating studies and extracting data.

A meta-analysis will assess the primary outcome of all-cause mortality at 90 days and secondary outcomes of ventilator and vasopressor free days, renal replacement therapy use, incidence of acute kidney injury, and patient quality of life outcomes.

The limitations of this review include the clinical heterogeneity of the included trials, diversity of the targeted population receiving fluid therapy, variability in the composition of balanced crystalloid solutions, and timing of initiation of study crystalloids. We will address all limitations with the Grading of Recommendations, Assessment, Development and Evaluations framework.

## Introduction

### Rationale

Selection of intravenous fluids for fluid therapy has been a perennial debate in the care of critically ill adults. With increasing data showing adverse outcomes from the use of hydroxyethyl starch (HES)^1,2^, and no significant advantage from the use of other colloid solutions,^3^ crystalloids have become the most commonly used fluids in critically ill patients.^4–6^ Although 0.9% sodium chloride (normal saline) has traditionally been the crystalloid of choice, concerns about its high chloride content and that its use might cause acute kidney injury (AKI) and/or increase the risk of death have resulted in increasing use of balanced salt solutions (BSS).^7,8^

The most robust trial data comparing BSS to normal saline in critically ill adults come from three cluster-randomised cluster-crossover trials (cRCTs). The 0.9% Saline vs. Plasma-Lyte 148 for ICU fluid Therapy (SPLIT)^9^ trial compared Plasma-Lyte 148 with normal saline in 2,278 adult critically ill patients. There was no difference in the primary outcome of AKI at 90 days. The isotonic Solution Administration Logistical Testing (SALT)^10^ pilot study compared BSS (Ringer’s solution or Plasma-Lyte A) to normal saline in 974 critically ill patients - the primary outcome related to feasibility, but reported no differences between groups in the secondary outcome of Major Adverse Kidney Events within 30 days (MAKE 30). The much larger Isotonic Solutions and Major Adverse Renal Events (SMART)^11^ trial compared BSS (Ringer’s solution or Plasma-Lyte A) to normal saline in 15,082 critically ill patients and reported a statistically significant reduction in the primary outcome of MAKE 30 in patients treated with BSS.

The most reliable totality of evidence on which clinicians can base the treatment of their patients come from systematic review and meta-analysis (SRMAs) of randomised control trials that includes substantial number of patients who experience the end point of interest. For comparisons of BSS and normal saline the most important outcomes of interest are mortality, AKI, and treatment with renal replacement therapy (RRT) with two recent SRMAs reporting these outcomes.^12,13^ In approximately 20,000 patients from a total of 9 RCTS/cRCTs of BSS compared with normal saline, there were approximately 2500 deaths, close to 2,900 patients with acute kidney injury, and almost 3000 treated with RRT.^12,13^

Two large blinded RCTs comparing BSS to normal saline for fluid resuscitation and all fluid therapy are due to be published in 2021; The Plasma-Lyte versUs Saline (PLUS) Study^14^ recruited over 5000 critically ill adult patients in Australia and New Zealand, and the Balanced Solutions in Intensive Care Study (BaSICS)^15^ which recruited 11,000 critically ill adult patients in Brazil. Although the total number of outcome events for these trials is not yet known they will add substantial new information to inform the debate over the best crystalloid fluid for critically ill patients. We will include these new data in a trial level systematic review and meta-analysis to evaluate the effect of choice of crystalloid fluid therapy on mortality, AKI and other patient-centered outcomes, in critically ill adult patients.

### Objective

The primary objective of this systematic review and meta-analysis is to determine whether the use of BSS compared with normal saline for fluid therapy is associated with improved all-cause 90-day mortality in adult patients who are critically ill. Key secondary objectives will include receipt of new RRT and incidence of AKI along with other patient-centered outcomes.

### Methods and analysis

We will conduct a systematic review of RCTs and cRCTs with meta-analysis and will report the findings following the recommendations of the Cochrane Collaboration and the Preferred Reporting Items for Systematic Review and Meta-Analysis (PRISMA) statement. This systematic review has been registered on International Prospective Register of Systematic Reviews (PROSPERO) CRD42021243399.

### Eligibility criteria

#### Inclusion criteria

We will include all randomised controlled trials or cluster randomised controlled trials^16^ that meet the following criteria:

##### Population

Adult patients managed in a critical care unit^17^ This may include patients who are cared for in the emergency department and operating theatre but then transferred/treated in the intensive care unit or high dependency unit.

##### Intervention

Fluid therapy with a balanced crystalloid solution (low chloride solutions) such as Hartmann’s, Plasma-Lyte, and Lactated Ringer’s.^18^

##### Comparator

Fluid therapy with normal saline (also known as: 0.9% sodium chloride, 0.9% saline, isotonic saline).

##### Outcomes

Mortality and any other pre-specified outcomes of interest

### Exclusion criteria

- Non-randomised trials
- Trials of non-critically ill patients
- Trials where there is concern about scientific misconduct^19^

### Fluid therapy definition

We will include all trials of fluid therapy. Fluid therapy will likely be defined differently in the included trials. The reviewers will screen for trials that define fluid therapy as fluid for resuscitation only and also fluids for resuscitation plus ‘maintenance’ (fluid to provide normal daily water and electrolyte intake).

### Search strategy

We will systematically search Medline (using PUBMED), EMBASE, Cochrane Central registry of clinical trials (CENTRAL) and other clinical trials registries for potentially eligible trials. The search strategy will include MESH terms and keywords for:

*Balanced crystalloid solutions OR balanced salt solutions OR buffered crystalloids OR lactated ringer’s OR Hartmann’s OR Plasma-Lyte OR crystalloid OR normal saline OR 0*.*9% sodium chloride OR isotonic saline OR saline*

AND

*We will use sensitive search filters to identify randomised clinical trials and cluster randomised controlled trials*

We will limit the search to human adult studies with no restrictions on language, publication date or publication status. In addition, we will search the reference lists of relevant primary and review articles, clinical trial registries and published abstracts, as well as contacting experts in this field.

### Study records

#### Selection process

We will screen studies with the aid of a reference management system^20^ with a minimum of two investigators independently screening all retrieved references for inclusion based on a review of the study title and abstract. We will solve disagreements by referral to a third reviewer. We will then assess full text of identified papers by two investigators using the same approach as above. We will assess eligibility using the above inclusion and exclusion criteria. We will undertake quality assessment and data extraction of the included articles. We will provide a PRISMA^21^ flow diagram in the final report.

#### Data collection

We will extract data from included studies using a standardised data collection form. We will attempt to contact the corresponding authors to obtain missing data. We will have access to aggregate level data for the PLUS and BASICS study prior to publication as agreed by the investigators. We will not make data publicly available until after the main studies are published. We will extract all data in duplicate, with disputes resolved by discussion or resort to a third independent reviewer. The extraction form will capture information on study characteristics, including study design, methods and participant characteristics. We will collect data on the intervention and comparison, along with the primary and secondary outcome data. We will then compare the resulting data, and disparities resolved through discussion between the two reviewers, or if required, a third reviewer.

### Outcomes

The pre-defined outcomes investigated in this systematic review will be:

#### Primary outcome

All-cause 90-day mortality. If 90-day mortality outcomes are not reported in a trial, we will use the time point closest to day 90 (before or beyond).

#### Secondary outcomes

Where available we will report the following: mortality at longest timepoint, ventilator-free days (to day 28), vasopressor-free days (to day 28), receipt of new renal replacement therapy, incidence of acute kidney injury as defined in the original trial, and quality of life and functional outcome.

### Risk of bias

Two review authors, with no affiliation with any of the included RCTs, will independently assess the risk of bias for each included study. We will use all publicly available reports of trials, including published trial protocols and statistical analysis plans to assess risk of bias. We will have early access to PLUS and BASICS prior to the final manuscripts being published to determine risk of bias. We will resolve disagreements by discussions involving a third independent assessor if needed. We will use the Cochrane Collaborations tool for assessment of risk of bias specific for RCTs and cRCTs.^22^

We will judge studies to be at “overall low risk of bias” if they are assessed as having low risk of bias in all the above domains. If none of the studies are low risk of bias in all domains, we will formulate a group of lower risk of bias, which have low risk of bias in at least the random sequence generation, allocation concealment, blinding of outcome assessment and blinding of participants and personnel. We will then meta-analyse trials with “overall low risk of bias” if possible or lower risk of bias if too few studies have an “overall low risk of bias”.

We will present a ‘risk of bias summary’ figure, depicting each risk of bias item for each included study (red high risk, green low risk, yellow unclear).

Our primary conclusions and the presentation in the ‘summary of findings’ table will be based on the results of our analyses of studies with overall low risk of bias. If none of the studies are low risk of bias in all domains, we will secondarily base our conclusions on studies with lower risk of bias.

### Assessment of reporting biases

If we include ten or more studies, we will use funnel plots to assess reporting bias. For dichotomous outcomes we will test asymmetry with the Harbord test, and for continuous outcomes we will use the regression asymmetry test and the adjusted rank correlation.

### Assessment of evidence (GRADE)

We will use the Grading of Recommendations Assessment, Development, and Evaluation (GRADE)^23^ approach to assess the overall certainty of evidence for each primary and secondary outcome measure and present the results in the ‘summary of findings’ table. The certainty of evidence and our confidence in the effect-estimates will be evaluated based on study design, study quality, precision, consistency, directness and the risk of reporting bias. Consequently, we will rate the overall certainty of evidence as “high”, “moderate”, “low” or “very low” for each outcome

### Statistical analysis

We will perform Hartung-Knapp-Sidik-Jonkman random-effects aggregate data meta-analysis whenever it is feasible to do so (upon completion of the extraction process we will perform a feasibility assessment to evaluate the appropriateness of pooling each outcome).^24^ We will also assess pooled effects according to other random-effects models (such as the Der-Simonian Laird) to evaluate the robustness of the estimates (and their uncertainty) according to different estimates of the between-study variance (τ^2^). To better handle the uncertainty of the various parameters to be estimated (especially the between-study variance) and to allow the size of the treatment effect to be described directly in probability terms we will also perform Bayesian Meta-Analysis. We will perform sensitivity analyses using different priors including vague and semi-informative priors.

For binary outcomes, we will use Risk Ratios (RR) with 95% confidence interval (CI) (or credible intervals, for the Bayesian Meta-Analysis), for continuous outcomes Mean Differences (MD) and for time-to-event data we will use hazard ratio (HR). We will assess quantitative heterogeneity by a formal test of homogeneity and evaluating the proportion of total variability due to heterogeneity rather than by sampling error (I^2^). We will assess small-study effects by regression-based Egger test and eyeball evaluation of the contour-enhanced funnel plots.

We will perform statistical analyses using Stata and R.

### Clustering effect

Cluster randomised trials are eligible for inclusion. If the studies did not account for the clustering effect, therefore leading to unit of analysis error, we will inflate appropriately the standard error as described previously.^25^

### Dealing with missing data

We will use the results based on analyses of the intention-to-treat populations, and we will try to obtain missing outcome data from the original study authors. Missing data will not be imputed.

### Predefined Subgroup analysis

We will only assess the primary outcome in the predefined subgroups with analyses considered hypothesis generating.

Trial level sub-populations:

- Trials of low risk of bias vs. high risk or unclear risk of bias
- Cluster randomised vs. individual patient randomised
- Type of BSS vs. 0.9% saline
- Maintenance fluids only vs. All fluid therapy (resuscitation fluid plus maintenance fluids)

We will include patient level diagnostic subgroups if enough baseline data are available:

- Sepsis vs. no sepsis
- Trauma vs. no trauma
- Traumatic Brain Injury vs. no traumatic brain injury
- Diabetic ketoacidosis (DKA) vs. no DKA
- Cardiac surgery vs. others

### Patient and Public Involvement

We did not involve patient or consumer representative in the development of this protocol.

### Ethics and dissemination

This review does not require ethical approval as this is a systematic review of published studies. The results of this systematic review will be presented at national and international scientific meetings, will be submitted to a peer reviewed journal for publication and made available on publicly accessible institutional websites. The results of the trial level meta-analysis will not be publicly released prior to the results of all individual trials being publicly available without the express consent of the component trial authors.

### Discussion and limitations

This systematic review and meta-analysis will provide the most up-to-date synthesis of evidence on the use of buffered crystalloid solutions compared with normal saline on mortality and other important patient centered outcomes in adult critically ill patients.

We acknowledge limitations to the proposed systematic review. Included studies are anticipated to be heterogeneous in nature due to variations in fluid therapy definitions, location of critically ill patients, composition of balanced crystalloid solutions, differences in mortality time points, and differences in secondary outcomes definitions. In addition, the strength of a systematic review and meta-analysis relies in part on the strength of available studies, and therefore may be limited due to the lack of large randomised controlled trials in this area.

## Data Availability

Results are available in published studies

## Contributions

NH is the guarantor of the study. All authors were responsible for the concept of the study. NH, SF and AD made substantial contributions to the concept and design of the study, drafting of the study protocol, substantial contributions to the analysis plan, revising the work critically for important intellectual content and providing approval for the final version to be published. GLD and FZ made substantial contributions to the trial design and analytical plan, revising the work critically for important intellectual content and provided approval for the final version to be published. All other authors made substantial contributions to the drafting of the study protocol, revised the work critically for important intellectual content and provided approval for the final version to be published

